# Health impact of nuclear waste water discharge from the Fukushima Daiichi nuclear plant

**DOI:** 10.1101/2023.04.23.23288984

**Authors:** Huipeng Liao, Danyi Sun, Zifan Yang, Wenyu Huang, Qian Di

## Abstract

Tokyo Electric Power Company announced to discharge the contaminated radioactive water resulting from the Fukushima nuclear accident into the ocean after purification from 2023. Concerns remain about safety and removal efficiency of radionuclides. This study calculated the total radioactivity and simulated the marine transport of ^137^Cs, ^90^Sr, and tritium. It assessed activity concentration in ocean and marine products, lifetime doses from marine product consumption, and associated cancer risks. We found the radionuclides would be globally distributed and penetrate into deep ocean, with the highest concentrations along Japan’s eastern coast. If ^137^Cs and ^90^Sr were not removed, related cancer risks would range between 8.64 – 33.35 cases per 100,000 people, depending on age and discharge scenario. Risks would be below one case per 100,000 if only tritium is present. Efficient removal of radionuclides is crucial to mitigating health risks. This study provides evidence of potential health risks and recommendations for prevention.

## INTRODUCTION

The Fukushima nuclear accident occurred on March 11, 2011, following a massive 9.0 magnitude earthquake^1^. A tsunami cause power outage and three reactors at the Fukushima Daiichi nuclear power plant to meltdown. The cooling system was destroyed, and water has been constantly injected into the damaged reactors for cooling down, resulting in a large amount of waste water contaminated by various radionuclides in high concentration and stored in storage tanks. This process resulted in a large amount of contaminated water containing radioactive isotopes. Although several options for handling tritiated water have been discussed, such as geosphere injection, underground burial, or hydrogen release^2^, Tokyo Electric Power Company (TEPCO) decided to discharge stored tritium water into the Pacific Ocean starting in 2023^3^.

Advanced Liquid Processing System (ALPS) was designed to filter radionuclides before discharging into ocean^4^. This system is expected remove most radionuclides below regulatory limit, except tritium, an isotope of hydrogen and a weak beta-emitter, exists in the contaminated water in the form of tritiated water^4^. TEPCO has committed to follow related safety regulations and discharge tritium water with an annual limit of 22 TBq^5^. One study also pointed out that tritiated water should be diluted to meet the regulation, which specify a legal discharge concentration of 0.06 MBq/L^4^. The announcement of contaminated water discharge led to opposition from local fishermen and neighboring countries as well as concerns in academia. To understand the impact of tritium, researchers estimated the spatial dispersion of tritiated water under several release scenario using 3D global transport model^6^, calculated tritium concentration along Fukushima coast, in North America, and East Asia^7^, estimated activity concentration in water, sediments and biota, and effective dose^8^, and projected economic impact and social welfare loss^9^. As a weak source of beta radiation, ionizing radiation from tritium cannot penetrate the skin but can increase cancer risk if tritium is consumed. However, few studies have estimated public health risks induced by contaminated water discharge.

Apart from tritium, ^137^Cs in the contaminated water and released in the nuclear accident also received attention. Due to released high-energy gamma ray, ^137^Cs can impose noticeable environmental and health risks. Thus, several studies have focused on the ^137^Cs release and related health risks from Fukushima accident: researchers have estimated ^137^Cs release during the accident, simulated its oceanic circulation and penetration into deep ocean^10^, estimated its concentration in seawater along coastal areas in Japan and the Pacific Ocean^11^, calculated its activity concentration in ocean sediments^12^, and assessed related cancer incidents^13^. Different from ^131^I with half-life 8 days, ^137^Cs has relative long half-lives of 30 years and can impose long-last impacts; ^137^Cs can also transport long distance due to its high solubility. All above factors make ^137^Cs another potential radionuclide of concern. ALPS is supposed to remove all non-tritium radionuclides, including ^137^Cs^4^. Although there has been wide concern over the efficiency and safety of ALPS, relevant scientific and technical literature on contaminated water treatment of ALPS is limited^4^. Even concerningly, Associated Press reported 24 out of 25 filters attached to water treatment were damaged in 2021^14^; Science News also reported varying concentrations of ^137^Cs and ^90^Sr, which may pose questions on the functioning of the filtration system^3^. It is important to understand the potential amount of other non-tritium radionuclides, such as ^137^Cs, and prepared for worse situations. However, few studies have estimated the total activity of radionuclides that *could have* been discharged into the ocean if the filtration system were to dysfunction.

Therefore, in this study, we aimed to estimate the environment impact and health risks of the contaminated water discharge under possible worst scenarios. According to the study framework in Fig.S1, we first calculated the type and maximal amount of radionuclides that could have been produced during the Fukushima nuclear accident and identified radionuclides that have greatest health risks. Second, we performed global 3-dimensional transport model to simulate the transport and distribution of radionuclides under different scenarios. Third, we estimated the effective doses for ingested radionuclides and associated cancer risk. This study can help us understand the environmental impact and health risks of contaminated water discharge that may happen, and encourage to take proper actions to minimize risks.

## METHODS

### Amount of radionuclide activity concentration

We calculated the type of radionuclides and radioactivity due to the nuclear accident, given the fact that there exist relationships between radioactivity with half-life, fission product and reactor power^15^. First, the relationship between radioactivity and half-life period was given as: *A* = 0.693 *N/T*_1/2_, where A was the radioactive activity; *N* was the number of radionuclides; *T*_1/2_ was a half-life period. Second, the fission products were estimated using the following ways: if the irradiation time was longer than the half-life of a fission product, the activity of the product could reach equilibrium: *A* = 310*YP*, where A was the activity (10^12^ Bq), Y was the fission yield (percentage) of the nuclide, and P was the reactor power (MW). This formula can be used to estimate the activity of important nuclides such as ^133^Xe and ^131^I. If the half-life of nuclide was significantly longer than the irradiation time, its activity would increase linearly with time: 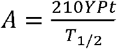 where *t* is the exposure time. Important radioactive fission products included ^131^I, ^134^Cs, ^137^Cs, ^90^Sr, ^106^Ru, and ^3^H (Table S1). Since activity concentration was proportion to the extent of damage of nuclear reactors, we considered several hypothetical scenarios and calculated corresponding damaged thermal power.

We considered in total three hypothetical conditions and calculated their upper limits of radioactivity, based on the information on status of in Fukushima nuclear power plants^16^. The total amount of radionuclides calculated according to both the damage condition of the core, and all radionuclides yielded from spent fuel was assumed to all enter the external environment. **For Condition 1**, we calculated damaged thermal power based on open reports, and total amount of radioactive substances based on damage condition of cores. In this case, the power of reactor No. 1 was 460 MWe and assumed a total meltdown. For No. 2 nuclear reactor, it was estimated a 35% damage of the reactor core of 784 MWe and hence a meltdown reactor core was 274.4 MWe [ref]. Reactor No. 3, which had a capacity of 784 MWe, was estimated to suffer 30% damage and had a meltdown reactor core of 235.2 MWe. For No. 4 nuclear reactor, there was no melting inside. Based on the above damage conditions, we calculated the corresponding total electrical power of 969.6 MWe. In addition, the spent fuel pools of four nuclear reactors, which generate heat (converted into electricity power for analysis) and radionuclides, were damaged or washed away. Furthermore, the electrical power of spent fuel pool of four nuclear reactors were 16.79 MWe, 41.99 MWe, 36.77 MWe and 95.21 MWe, respectively. The corresponding electricity power of the spent fuel pool was 190.76 MWe. In total, the total electric power influenced by the melting reactor cores and spent fuel pools under Condition 1 is 1,160 MWe. Considering that the thermal efficiency of the nuclear reactors was 33%, the thermal power is about 3,500 MWt (thermal power in MW). **For Condition 2**, all four reactor cores were assumed to generate radionuclides with full percentages and all radioactive materials to enter the external environment. The total electric power under condition 3 was 2,800 MWe. Taking into account of the thermal efficiency 33%, the corresponding thermal power was about 8,400 MWt. **For Condition 3**, we assumed a hypothetical damaged condition between Condition 1 and 2. We assumed the total damaged electrical power to be 1,600 MWe, and thermal power 4,800 MWt, given thermal efficiency of 33%.

### Radionuclides transportation simulation

Ocean transportation simulation was performed using Nucleus for European Modeling of the Ocean (NEMO) general ocean model version 4.0 with the Tracers in Ocean Paradigm (TOP) version^17^. We performed 50-year simulations of oceanic transportation with 2° × 2° horizontal grid and 20-year simulation with 0.25° × 0.25° horizontal grid to obtain high-resolution regional distribution of radionuclides. Vertical depth started from the ocean surface to the bottom of 5250 meters over 31 and 46 levels for the coarse-grid and fine-grid simulations, respectively. Considering total activity concentration and half-life, our global oceanic focused on the transportation of ^137^Cs, ^90^Sr, and tritium, with total released activity concentration from Condition 3. We designed 4 discharge scenarios where all radionuclides were discharged into the ocean within 1 month (Scenario 1), 1 year (Scenario 2), 5 years (Scenario 3), and 30 years (Scenario 4). We extracted monthly averaged activity concentration of ^137^Cs, ^90^Sr, and tritium, analyzed their spatial and vertical distribution, and performed follow-up health risk assessment. We also estimated the spatial coverage of influenced area due to contaminated water discharge, and defined influenced area where the concentration increased due to released ^3^H, ^137^Cs, and ^90^Sr above 0.001 Bq/m^3^.

### Exposure and risk assessments

After discharging, nuclear radiation from contaminated water could impose health risks if people consume contaminated aquatic organisms. To estimate health risks associated with radioactive water, we first estimated radioactivity increase in marine products due to contaminated water discharge. Aquatic organisms reach equilibrium of radionuclides with surrounding water column^18^. Although such equilibrium assumption may not hold in the short term and biokinetic model should be required to take dynamic process into consideration^19^, we still made equilibrium assumption in this study considering the long time scale of our study. The ratio of activity concentration between organisms and background environment under equilibrium could be used to predict activity concentration in aquatic organisms. According to International Atomic Energy Agency (IAEA), concentration factors for ^137^Cs and ^90^Sr were 100 and 3, respectively^20^, and we calculated radioactivity retained by the aquatic organism based on the concentration factor and activity concentration of surrounding water column^19^. For tritium, although previous research observed high concentration factor between sea water and aquatic organism^21^, we still used the concentration factor of tritium of 1 from IAEA^20^. We considered radionuclide uptake by marine organisms from the surface level to around 200-meter deep, the depth where most ocean fishery takes place. We extracted activity concentration of radionuclides from simulation outputs, and predicted activity concentration of marine organisms based on concentration factors. Above calculation process was made for every grid cell and by year to capture spatiotemporal variation in radionuclides.

The amount of marine product intake was obtained from Japan’s National Health and Nutrition Survey in 2019^22^. Amount of marine products consumption was estimated for different age groups, including 3-month old (0-1 years of age), 1 year (1 – 2 years of age), 5 year (3 – 7 years of age), 10 year (8 – 12 years of age), 15 year (13 - 17 years of age), and adults (above 18 years of age), based on International Commission on Radiological Protection (ICRP) report^23^. We also used the same age groupings for dose calculation.

Next, we estimated effective dose on a group of hypothetical persons of different age groups starting from 2023, who will be exposed to radionuclides due to consuming contaminated marine products captured near Fukushima. For a particular year *i*, the committed effective dose (mSv/year) of a particular radionuclide *j* due to ingesting contaminated marine products was calculated as the product of consumed marine products per year (*M*, kg), radioactivity level of radionuclide *j* in marine products (*C*, Bq/kg), and age-dependent committed effective dose coefficient from ingestion of radionuclide *j* (*e*_*j*_, mSv/Bq) in the following equation. Effective dose coefficients were obtained from ICRP reports^23^. Total committed effective dose were the sum of committed effective dose by different years over the study period^24^.

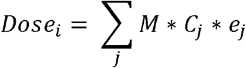

Finally, we estimated associated health risks and considered all solid cancers, thyroid cancer, and leukemia as health outcomes of interests for their radiosensitivity. The lifetime nominal risk was estimated by multiplying the total committed effective dose and adjusted nominal risk coefficient. We considered lifetime nominal risk from the age of first exposure (i.e., the start of contaminated water discharge) until the end of 50-year simulation, whichever comes first. According to ICRP reports, the adjusted nonmail risk coefficients for solid cancers, thyroid cancer, and leukemia were 4.871%/Sv, 0.127%/Sv, and 0.615%/Sv, respectively^25^.

Above exposure and risk assessment processes were repeated for all 4 discharge scenarios; as a comparison, we also designed a fifth scenario (Scenario 5) and calculated associated exposure and cancer risks. Scenario 5 was based on Scenario 4 with only tritium, i.e., all contaminated water completed discharge 30 years and all non-tritium radionuclides were removed.

## RESULTS

### Total amount of radionuclides

The total initial activity concentration of radionuclides produced by nuclear accident were 5,370 PBq (Condition 1), 12,900 PBq (Condition 2), and 7,370 PBq (Condition 3), respectively (Table S2, Table S3, and Table 1). The initial activity concentration of the three scenarios were somehow similar and at the same magnitude. We chose Condition 3 for the following analysis, since Condition 3 was a reasonable balance between Condition 1 and 2.

**Table 1.** Total amount of radionuclides after Fukushima Daiichi nuclear power plant accident.

| Radionuclides | Half-life (a) | Activity concentration per unit (10<sup>12</sup> Bq /MW) | Initial activity concentration (10<sup>12</sup> Bq) | Activity concentration after 12 years (10<sup>12</sup> Bq) |
| --- | --- | --- | --- | --- |
| <sup>3</sup>H | 12.3000 | 1.4200 | 6816.0000 | 3466.6033 |
| <sup>131</sup>I | 0.0222 | 940.0000 | 4,512,000.0000 | - |
| <sup>134</sup>Cs | 2.1000 | 140.0000 | 672,000.0000 | 12,810.4100 |
| <sup>137</sup>Cs | 30.1000 | 70.0000 | 336,000.0000 | 254,8900 |
| <sup>90</sup>Sr | 28.9000 | 52.0000 | 249,600.0000 | 189,520.2000 |
| <sup>106</sup>Ru | 1.0000 | 310.0000 | 1,488,000.0000 | 363.9234 |
| <sup>238</sup>Pu | 89.0000 | 1.3000 | 6,240.0000 | 5,683.3560 |
| <sup>239</sup>Pu | 24,000.0000 | 0.2800 | 1,344.0000 | 1,343.5340 |
| <sup>240</sup>Pu | 6,580.0000 | 0.3100 | 1,488.0000 | 1,486.1210 |
| <sup>241</sup>Pu | 14.7000 | 5.6000 | 26,880.0000 | 15,266.6100 |
| <sup>242</sup>Pu | 380,000.000<br>0 | 0.0005 | 2.4000 | 2.39995 |
| <sup>242</sup>Cm | 0.4500 | 15.0000 | 72,000.0000 | 0.0007 |
| <sup>244</sup>Cm | 18.2000 | 0.9100 | 4,368.0000 | 2,765.9440 |

Note: For this condition (Condition 3), we assumed a hypothetical damaged condition with radioactive substance amount between those in Condition 1 and 2. More details were articulated in the Method Section. <sup>131</sup>I would be rarely found after 12 years of decay.

For Condition 3, among these radionuclides, ^131^I contributed the highest proportion of initial radioactivity and the upper limit of total radioactivity level reached 4,510 PBq (Table 1), and 940 PBq equivalent ^131^I had been released after the accident according to previous reports^26^; as a comparison, radioactive release from the Chernobyl incident was about 6 times larger^27^ ^131^I was not considered in the follow-up analysis for its short half-life of 8 days. We also excluded ^106^Ru from the follow-up analysis also for its short half-life. Tritium was of interest, since tritium in contaminated water received much public attention. For tritium, the half-life of tritium is 12.3a and the unit power activity is 1.42^*^10^12^Bq/MW; we calculated upper amounts of tritium under different conditions and these values were of the same magnitude of the tritium amount revealed from open reports and studies^6,28,29^ (Table 1, Table S2, Table S3).

There were two radionuclides deserving greater attention: ^137^Cs and ^90^Sr had relatively high initial activity concentrations of 336 PBq and 250 PBq, respectively. The two radionuclides also had much longer half-lives of 30.1 and 28.9 years, respectively. Previous studies on environmental radioactivity monitoring also chose the two radionuclides as radionuclides of interests^30-32^. Thus, we focused on ^137^Cs, ^90^Sr, and ^3^H for the following simulations and health analyses.

### Spatial distribution of radionuclides

Video S1-S3 revealed the spatial dynamics of ^3^H, ^137^Cs, and ^90^Sr under four discharge scenarios over the next 50 years, and Fig.1 displays the same dynamics after 1, 5, 15, and 50 years. Under all scenarios, released ^3^H, ^137^Cs, and ^90^Sr would be highly concentrated in oceans surrounding Japan for the first year; and hotspots of radionuclides moved eastwards along ocean current and covered the Northern Pacific Ocean after 5 years. Five years after radioactive water release, for Scenario 1 and 2 under which contaminated water completed discharge within 1 month and 1 year, respective, the activity concentration near Fukushima decreased noticeably. Nonetheless, released contaminated water could reach most of areas in the Pacific and Indian Oceans, appear in the west coast of North America, even cross Bering Strait and enter the Arctic Ocean, 15 years after discharge initiation; the similar pattern also held for Scenario 3 and 4. Under all scenarios, ^137^Cs and ^90^Sr could also eventually spread over the oceans, demonstrating a similar spatial pattern; ^3^H would spread globally but not the surrounding water of Antarctica.

**Fig. 1.**
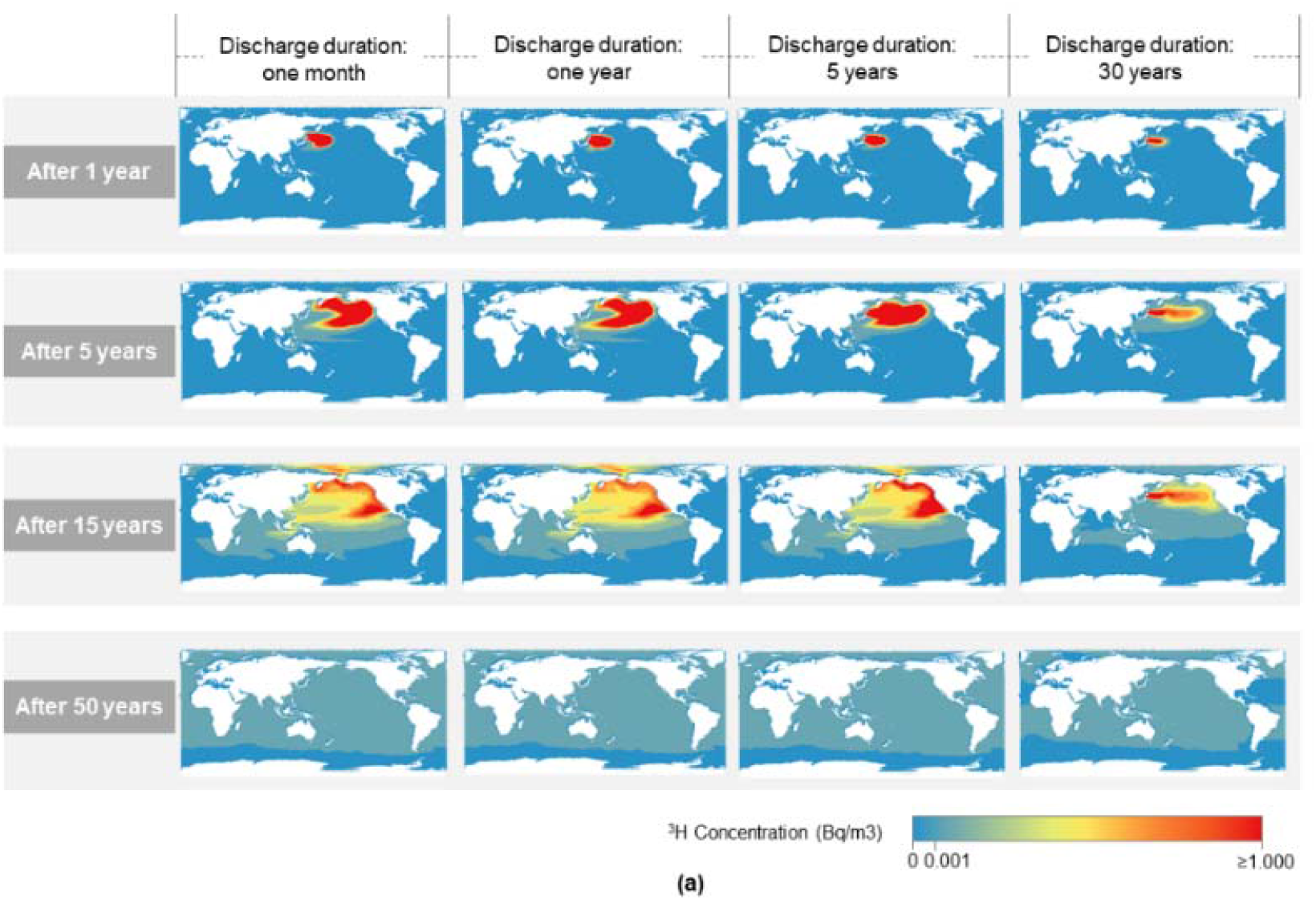
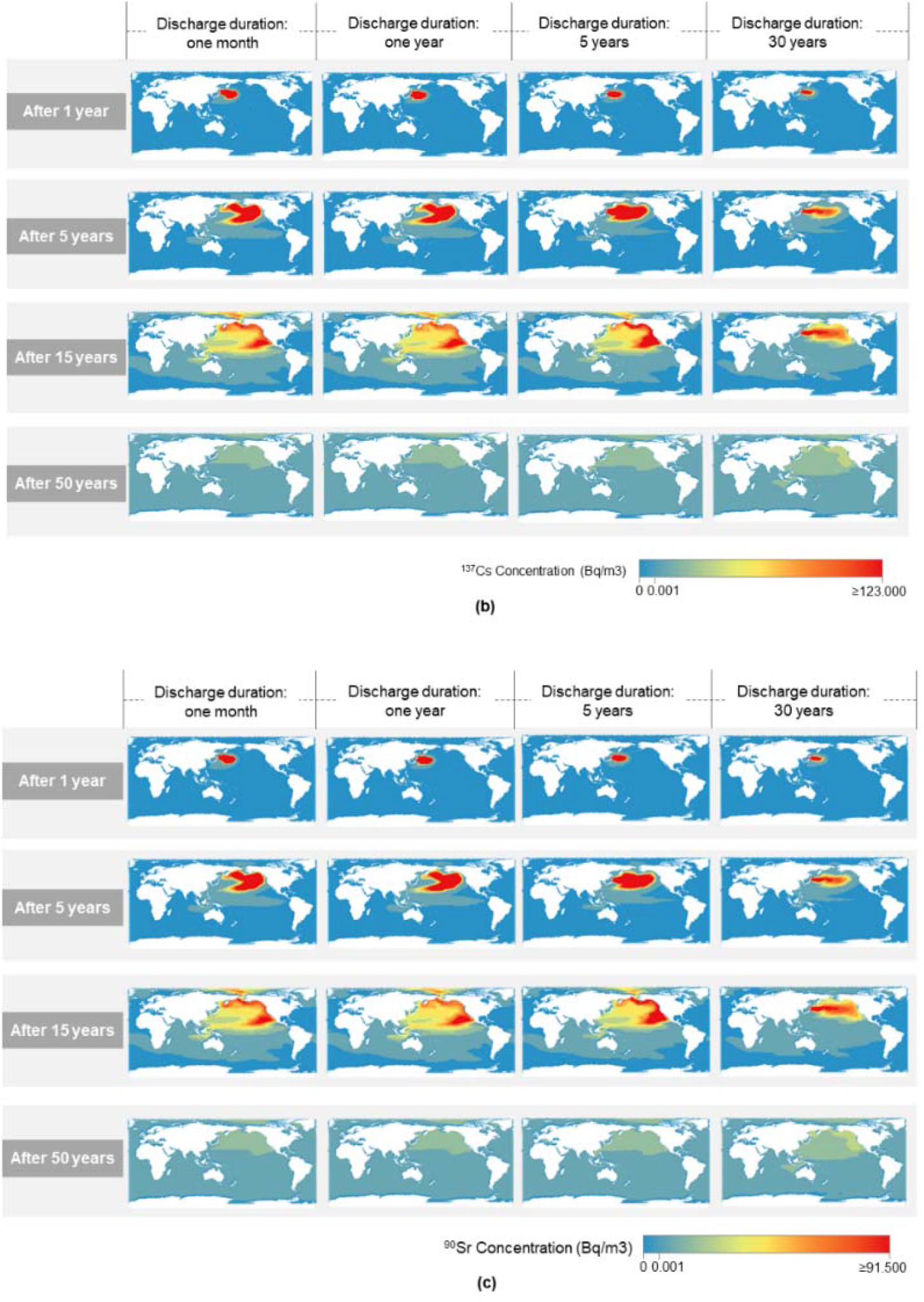
Spatial distribution of activity concentration by radionuclides under different discharge scenarios. (a) Spatial distribution of ^3^H concentration if it was discharged into the ocean within one month, one year, 5 years, or 30 years; (b) Spatial distribution of ^137^Cs concentration if it was discharged into the ocean within one month, one year, 5 years, or 30 years; (c) Spatial distribution of ^90^Sr concentration if it was discharged into the ocean within one month, one year, 5 years, or 30 years.

We next estimated the spatial coverage of influenced area due to contaminated water discharge. The estimated influenced area differed by radionuclide types, discharge duration, and years after discharge initiation (Fig.2). Overall, a longer discharge duration was associated with a slower increase in the influenced area. Under Scenarios 1, 2, and 3, where the discharge duration was 1 month, 1 year, or 5 years, respectively, the influenced area due to released ^3^H, ^137^Cs, and ^90^Sr were expected to spread over 77% after 40-45 years, over 99% after 36 years, and over 99% after 36-39 years, respectively. When the release was completed in 30 years (i.e., Scenario 4), the spread of radionuclides could be slower compared to other discharge scenarios, with the influenced area gradually increasing to 65% for ^3^H, 99% for ^137^Cs, and 99% for ^90^Sr after 49-50 years of discharge. Taking together, ^137^Cs and ^90^Sr would be found globally and eventually spread to almost all oceans, while ^3^H would also spread widely, although to a lesser extent.

**Fig. 2.**
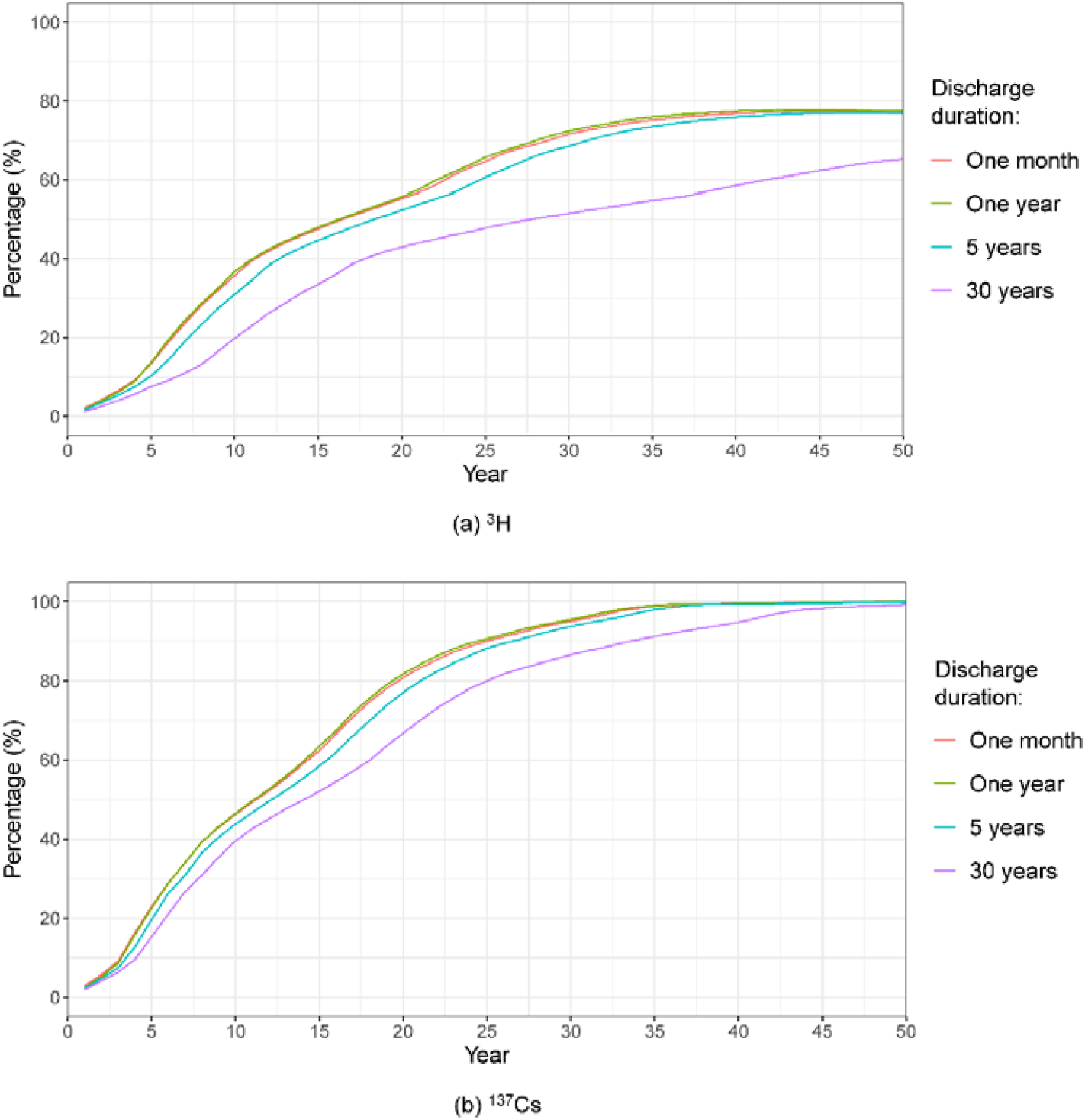
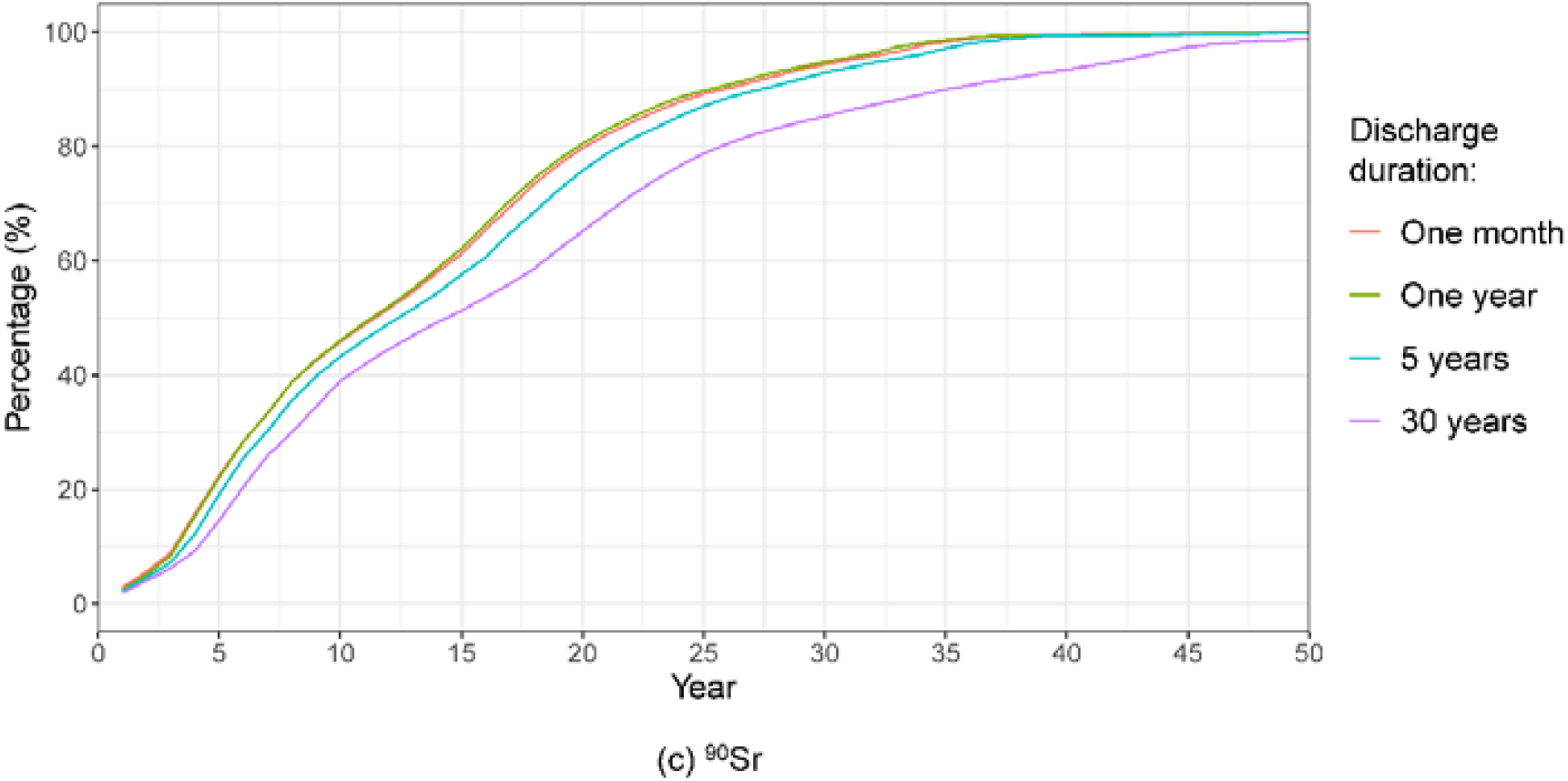
Influenced areas due to contaminated water discharge under different discharge scenarios. (a) ^3^H-influenced areas due to contaminated water discharge if the water was released in one month, one year, 5 years, or 30 years; (b) ^137^Cs-influenced areas due to contaminated water discharge if the water was released in one month, one year, 5 years, or 30 years; (c) ^90^Sr-influenced areas due to contaminated water discharge if the water was released in one month, one year, 5 years, or 30 years. Influenced area was defined where the activity concentration increases due to released ^3^H, ^137^Cs, and ^90^Sr was above 0.001 Bq/m^3^.

### Detailed distributions in regions of interests

Patterns of ^3^H, ^137^Cs, and ^90^Sr differ by discharge scenarios and geographic locations. We chose coasts of Fukushima, Yangtze River Delta (China), Papua New Guinea, and Hawaii (United States) as location of interests (Fig.3) to investigate the distribution of radionuclides with high resolution. In general, ^3^H, ^137^Cs, and ^90^Sr were highly similar in time trend but different in terms of peak concentrations. For example, when the contaminated water discharge was completed in one month (i.e., Scenario 1), Fukushima could witness peaks of all radionuclides in its adjacent sea area within the first year, with peak concentrations of ^3^H 421.92 Bq/m^3, 137^Cs 31,182.33 Bq/m^3^, and ^90^Sr 23,185.41 Bq/m^3^, with orders of magnitudes of difference. The highest value of activity concentration and the associated peak time in its adjacent sea area depend on the geographic location. Under all discharge scenarios, oceans surrounding Fukushima received the highest peak value of activity concentration, followed by that near Hawaii, Papua New Guinea, and Yangtze River Delta; the activity concentration of sea water near Fukushima reached peak value first, followed by that near Hawaii, Yangtze River Delta, and Papua New Guinea.

**Fig. 3.**
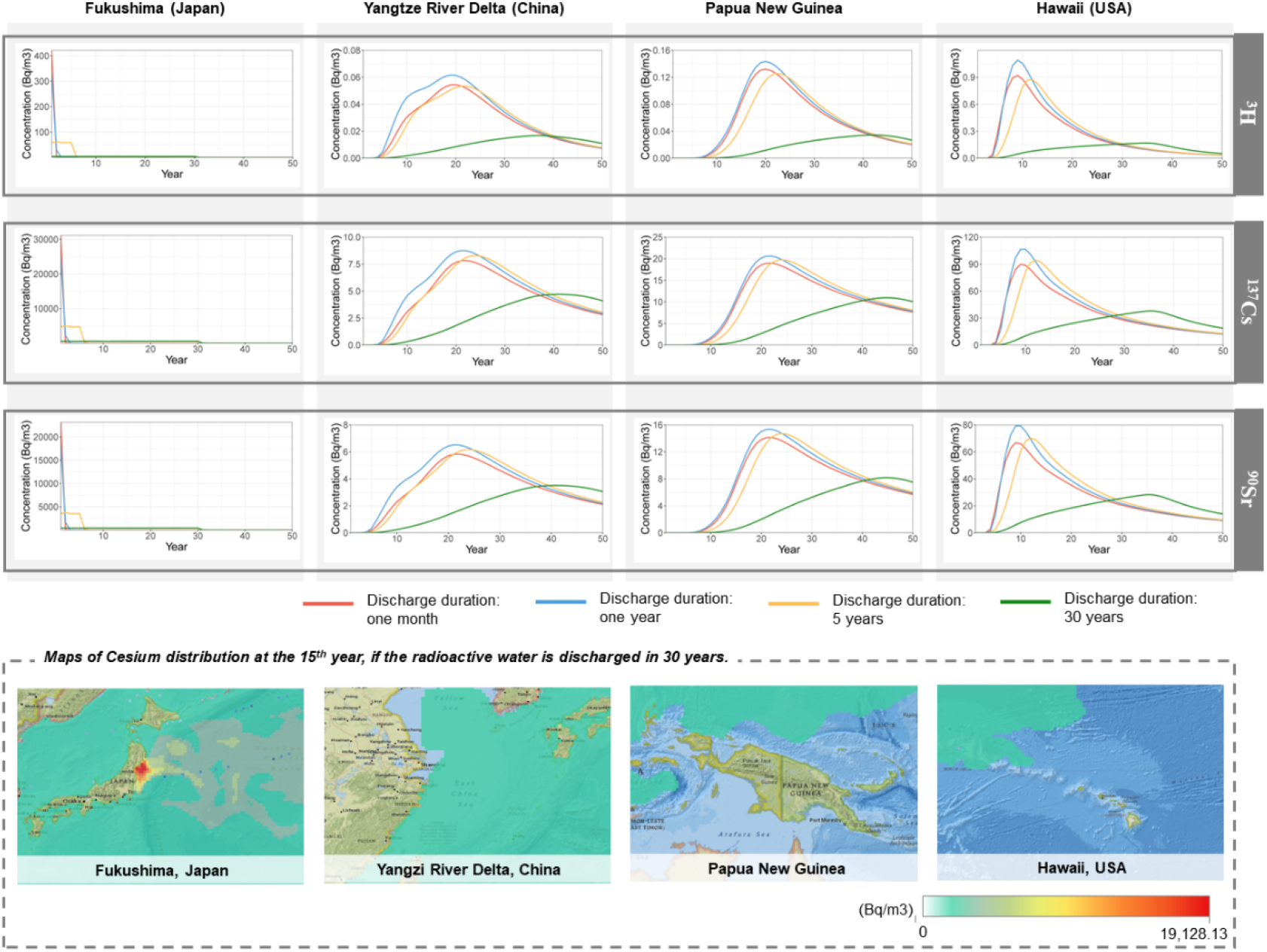
Activity concentrations of sea surface water in Fukushima (Japan), Yangtze River Delta (China), Papua New Guinea, and Hawaii (USA) by radionuclides and release scenarios.

In addition, for a particular location, the peak value and peak time of activity concentration were determined by the discharge plan. If the contaminated water discharge was completed within 1 month, 1 year, or 5 years, the activity concentration of a radionuclide would peak at a similar time and value. If the discharge was completed in 30 years, the peak would delay with a lower value. For example, ^137^Cs concentration near Papua New Guinea would meet the maximum in the range of 18-21 Bq/m^3^ after 22-24 years if the discharge duration was 1 month, 1 year, and 5 years (i.e., Scenarios 1-3). In the scenario where the discharge duration was 30 years (i.e., Scenario 4), ^137^Cs concentration near Papua New Guinea would peak 45 years after discharge initiation and reached the highest level of 10.98 Bq/m^3^, with delayed peak time and reduced value compare to other scenarios.

### Vertical distribution

We analyzed the vertical distribution of released radionuclides at 37.5°N - 38.8°N (Fig.4). Vertical distributions of ^3^H, ^137^Cs, and ^90^Sr were similar under all discharge scenarios. If contaminated water discharge was completed within 1 month (Scenario 1), most released ^3^H, ^137^Cs, and ^90^Sr would be concentrated on the longitude near 160°E and between depths of 0-217 meters for the first year, forming a ‘belt’ pattern. Such belt of contaminated water then moved eastwards with halved concentration in the 2^nd^ and 5^th^ year. Unlike Scenario 1, for Scenarios 2, 3, and 4 where corresponding discharge duration was 1 year, 5 years, and 30 years, the released ^3^H, ^137^Cs, and ^90^Sr were highly concentrated near Fukushima between the depths of around 0-106 meters for the first year. Overall, concerningly, contaminated water was able to penetrate into deep ocean rather than just surface water.

**Fig. 4.**
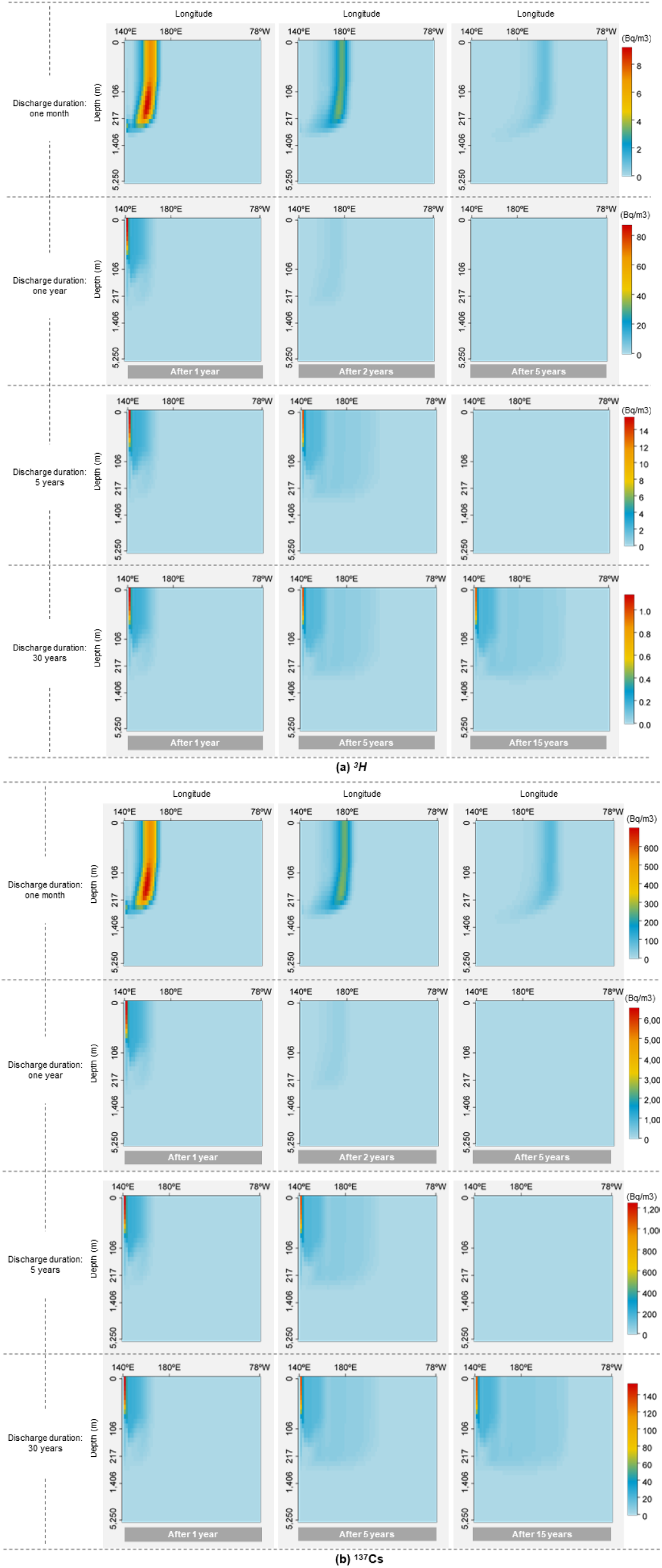
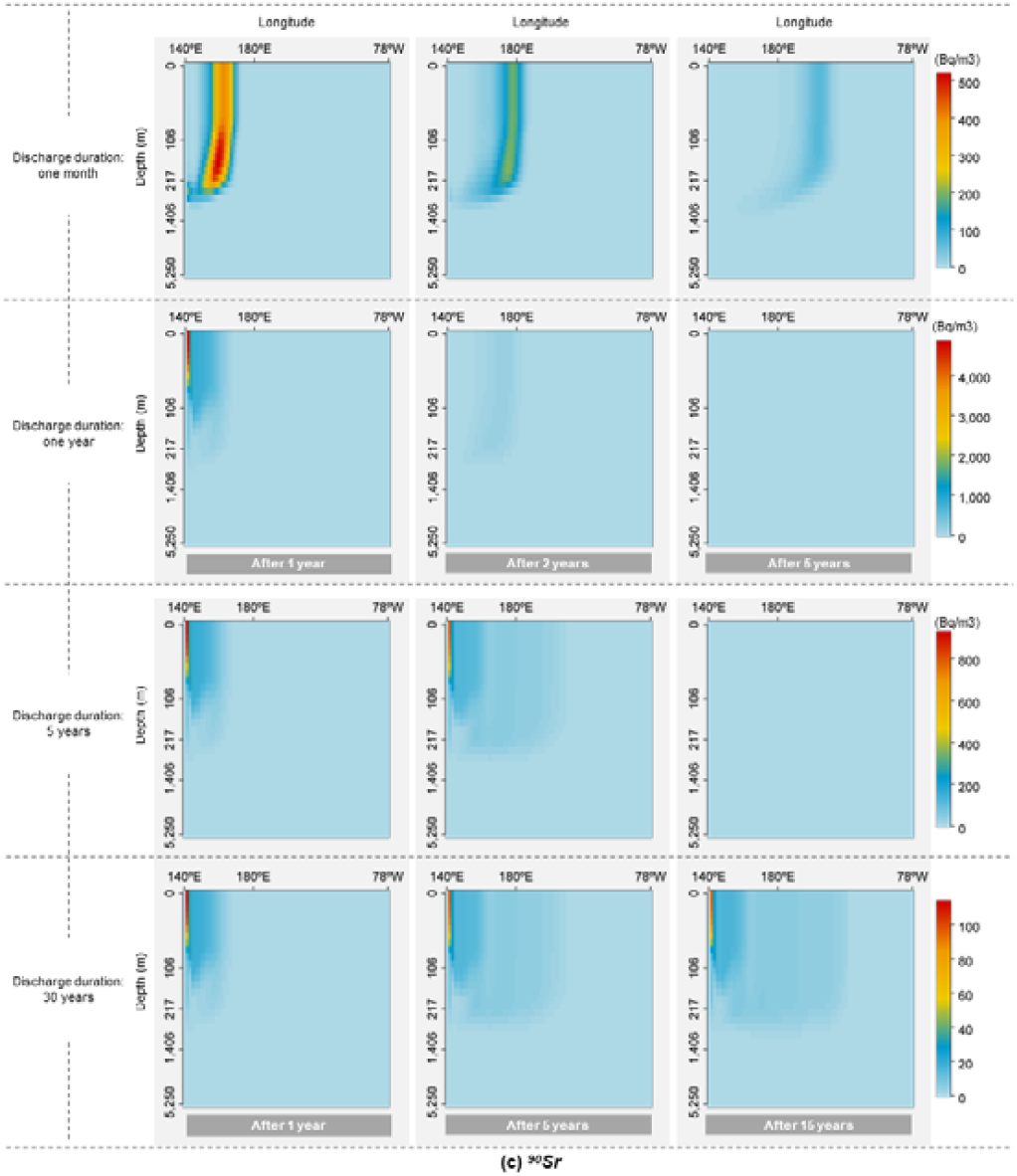
Vertical distribution of activity concentration at 37.5°N - 38.8°N by radionuclides under different discharge scenarios. (a) Vertical distribution of ^3^H concentration at 37.5°N - 38.8°N if contaminated water was discharged within one month, one year, 5 years, or 30 years; (b) Vertical distribution of ^137^Cs concentration at 37.5°N - 38.8°N if contaminated water was discharged within one month, one year, 5 years, or 30 years; (c) Vertical distribution of ^90^Sr concentration at 37.5°N - 38.8°N if contaminated water was discharged within one month, one year, 5 years, or 30 years.

### Cancer risks

Above simulations indicated coastal areas of Japan to be hotspot of possible radionuclide contamination with noticeable increase in activity concentration in sea water, while radioactivity increases in other parts of world was at smaller scale compared with the background level. Thus, we focused on the health risks due to consuming marine products captured in the vicinity of hotspots, i.e., coastal areas of Japan. We obtained amounts of marine products consumption from government statistics, focused on fish and shellfish consumption^22^, and calculated the lifetime doses for each age group between 2.2 to 6.8 mSv. The lifetime nominal risks for all solid cancers were magnitudes higher than risks of thyroid cancer and leukemia. The lifetime nominal risk of solid cancers ranged from 8.64 – 33.35 cases per 100,000 people. If only tritium was present in the contaminated water, associated cancer risk was below 1 case per 100,000 people for all age groups (Table 2).

**Table 2.**
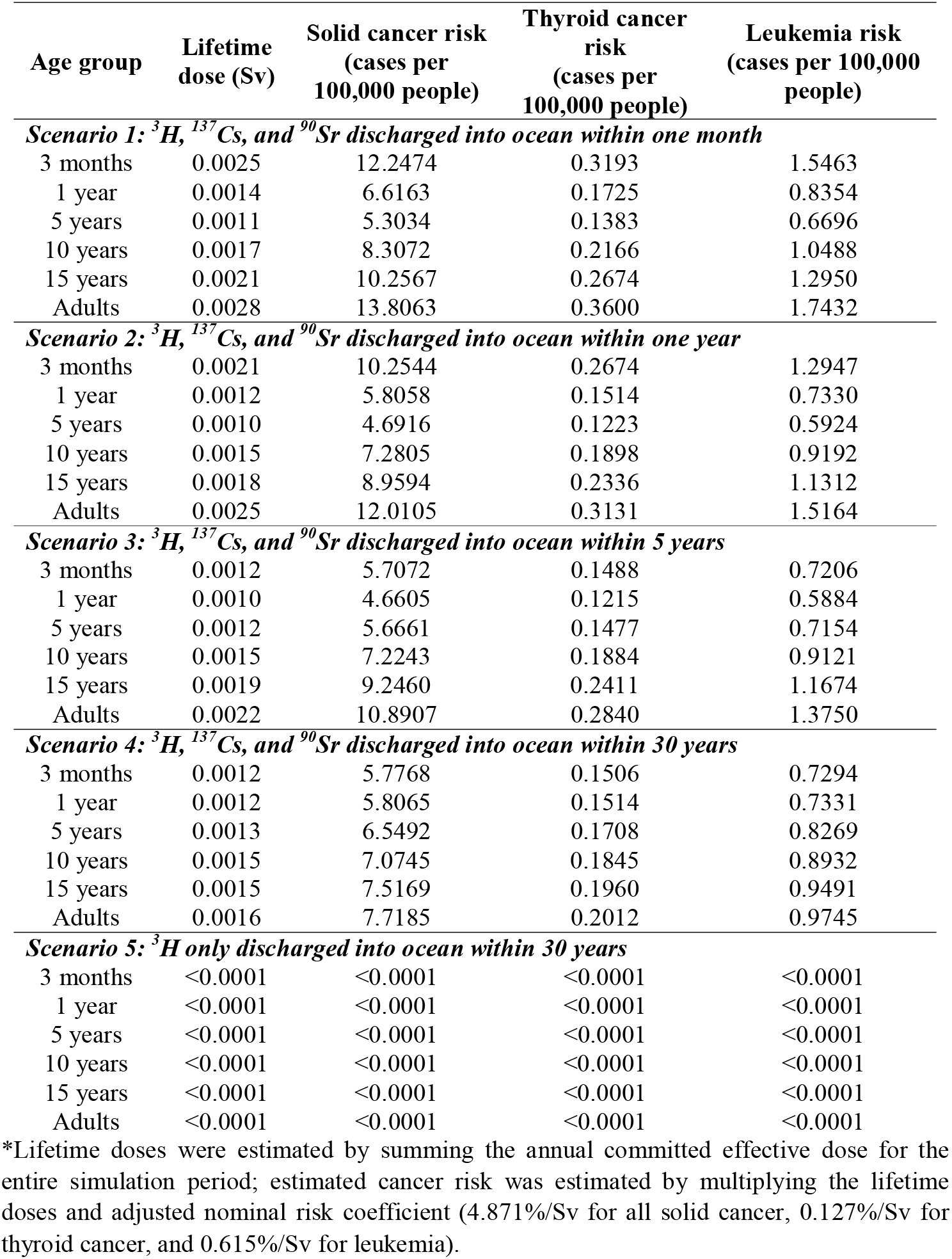
Cancer risk from dietary fish intake by age under different discharge scenarios.

## DISCUSSION

In this study, we first estimated the upper limits of radionuclides amount produced from Fukushima accidents, based on the damaged condition of each nuclear reactor. ^137^Cs, ^90^Sr and ^3^H were identified as radionuclides of interests. Several hypothetical scenarios were designed assuming different release durations and removal efficiencies of radionuclides. Global ocean circulation model simulated the transportation of radioactive contaminated water in a global scale. Simulation output found that decades after discharge, radionuclides from contaminated water will be found globally, even in faraway places such as Pacific and Indian Oceans. In particular, Arctic Ocean and Northern Polar area will also be affected, which has not been previously reported. Under all scenarios, Japanese Eastern coast will witness the highest increase of radiation level, with ^137^Cs radiation level increasing substantially compared with the background level. More concerningly, Northeastern Pacific Ocean is also the places contributes significant proportion of global marine fisheries catches (FAO citation). Based on our calculation, risks for all solid cancers ranged from 8.64 – 33.35 cases per 100,000 people if non-tritium radionuclides were not removed. Thus, the removal efficiency of non-tritium radionuclides is a critical factor determining the doses and cancer risks. Special efforts should be made to ensure the proper functioning of filtering system.

Radiocesium produced from Fukushima accident is a concern. Several studies estimated the inventory ^137^Cs from the damaged fuel between 760 – 820 PBq^33^, which is comparable to the 336 PBq estimated in our study. Among all produced ^137^Cs, about 4% were released into the atmosphere, and 16% of ^137^Cs inventory was released into the ocean when nuclear reactors were overflowed^34^. In terms of absolute amount, between 10 – 37 PBq of ^137^Cs was released into the atmosphere during the accident, although great difference existed between studies and the amount can differ by one magnitude^34^. Among ^137^Cs released into atmosphere, about 5.6 PBq of ^137^Cs was deposited back to Japan and the surrounding ocean; eastern and northeastern Japan were strongly contaminated but with noticeable heterogeneities^35^. Thus, it is crucial to understand the impact of radiocesium. However, to the best of our knowledge, few studies estimated the possible impact of radiocesium discharged into the ocean, since ALPS is assumed to remove radiocesium from the contaminated water. However, recent news cast doubts on the proper functioning and removal efficiency of ALPS^14^. Thus, it is important to calculate possible ^137^Cs amount in the contaminated water to be discharged into the ocean, and estimate the worst condition, i.e., the upper limit amount.

By assuming the worst condition, global transport model simulated the highest activity concentrations of ^137^Cs and other radionuclides in different geographic locations. Based on our 3D oceanic circulation model, the North Pacific would be mostly affected by Fukushima-related ^3^H, ^137^Cs, and ^90^Sr. Regions near or in the Pacific Ocean to the east of Japan, such as Hawaii, would witness relatively early peaks than other places. It is suggested to perform early monitoring on environmental and health impacts in these areas. Our simulation also indicated that the vertical distribution of ^3^H, ^137^Cs, and ^90^Sr can penetrate as deep as 106-217 meters in sea water near Fukushima, and similar results were also previously reported by a field study where Fukushima-related ^90^Sr was detected in the depth of 100-200 meters near Fukushima^36^. Putting our simulations and previous studies together, it is recommended to perform long-term surveillance for the sea surface as well as for a variety of depths, such as 0-250 meters.

Apart from ^137^Cs, ^3^H is another nuclear radionuclide existed in the contaminated water. Different from ^137^Cs, ^3^H will not be removed by ALPS. For the total amount of ^3^H, our estimation was similar to the open report made by TEPCO. Our 3D oceanic circulation model revealed the global transportation of ^3^H and other possible radionuclides in the contaminated water. Under the worst scenario, if there is a leakage and a sudden overflow of contaminated water into the ocean, ^3^H concentration along the Eastern coast of Japan will sharply increase by over 400 Bq/m^3^ and quickly decline afterwards. Considering a background concentration of ^3^H around 0.28 – 1.20 Bq/L^37^, the worst scenario will cause a noticeable increase of ^3^H level along Japanese coast. This contaminated water will flow eastwards and finally cause a mild ^3^H concentration increase of 2 Bq/m^3^ 10 years later along the Western coast of the United States. Under a stringent scenario (i.e., Scenario 4), however, the ^3^H increase along Eastern coast of Japan will be around 4 Bq/m^3^. We also simulated ^3^H increase using ^3^H data reported by TEPCO and obtained similar results. Thus, it is important to avoid the worst scenario where all or most stored contaminated water discharge into the ocean during a short period of time.

We found that potential cancer risks would be primarily due to ^137^Cs. Different from Sr and ^3^H, (1) Cs can form soluble compounds that facilitate its long-range transportation; (2) Cs can could retain in forms of mineral-free and particulate organic matter, and could remain potentially mobile and bio-available, imposing long-term environmental hazards^38^; (3) Cs also had concentration factors which were magnitude higher than other radionuclides; (4) ^137^Cs had high initial activity concentration and relative long half-life, with enough radioactivity remaining by the time contaminated water began to discharge. All above factor render ^137^Cs the radionuclide with greatest potential health risks. Previous study estimated health risks of ^137^Cs released from the Fukushima accident among residents living in contaminated villages near Fukushima nuclear power plant and found that internal doses from dietary intake and inhalation of ^137^Cs ranged from 0.0058 – 0.019 mSv/y and 0.31 – 0.53 mSv/y, respectively^39^. Dose due to dietary intake was at smaller scale, and this is expected, since inhalation pathway was to main contributor of dose in the Fukushima accident according to a World Heath Organization report^40^. If considering other radionuclides released into the atmosphere during the accident, there would be 130 cancer-related mortalities and 180 cancer-related morbidities^41^. Few studies have ever estimated health risks due to contaminated water discharge, and to the best of our knowledge, this is the first study on this topic. We found that lifetime doses for people who would regularly consume contaminated marine product ranged 2.2 to 6.8 mSv under the worst scenarios where non-tritium radionuclides were not removed. These doses were smaller compared with doses that people in European countries were exposed after Chernobyl accident^42^. It is worth mentioning that potential cancer risks were primarily due to ^137^Cs and consuming contaminated marine products captured from oceans surrounding Fukushima. Removing ^137^Cs efficiently from contaminated water or avoiding consuming contaminated fish should alleviate such cancer risks. While our study only assessed the environmental and health impact of TEPCO’s plans to release radioactive water into the Pacific Ocean, the plan is controversial with regards to international law. Multiple conventions, including convention on Nuclear Safety (CNS), the Convention on Early Notification, the London Dumping Convention and its Protocol, and the United Nations Convention on the Law of the Sea (UNCLOS), do provide sufficient constraints on Japan’s decision to discharge radioactive wastewater into the ocean^43^. However, it is not explicitly stated whether or not it is legal according to international law.

This study is among the first to assess health effect of contaminated waste water discharge from Fukushima accident and thus bears several limitations. First, the calculation failed to consider: (1) different types of marine products, (2) population growth, age structure of the population, change of fishery amount, changing preference of fish; (3) international trade of marine products; (3) the dynamic process of aquatic organism absorbing radionuclides but assumed equilibrium between fish and water column. Nevertheless, since the relationship between dose and cancer risk is a non-threshold linear one, putting above-mentioned issues into consideration only changes our results in a linear way. The exact number will differ, but the overall conclusion remains unchanged. Second, we mostly focus on cancer risk due to intake, ignoring other pathways such as dermal exposure. Besides, we did not consider health risk via intake of desalinated sea water; and it is expected that people will increasingly rely on desalinated sea water for drinking, irrigation, and other purposes in the future. Since sea desalination equipment is not commonly installed around Pacific region, we did not consider it in this study, but it deserves investigation in future studies. Third, parameters from ICRP, such as ingestion coefficients and nominal risk coefficients, bear uncertainty. This study also has several advantages. First, this study adopted an interdisciplinary approach and integrated nuclear physical, ocean dynamics, and public health to investigate the worst scenario of Fukushima nuclear waste water discharge and identified the determining factor. Second, different from other studies that focused on status quo, this study focused on the hypothetical worst scenario. This study can provide scientific evidence for responsible party and stakeholders on the worst scenarios and avoid the worst scenarios to protect health of all humans.

## Data Availability

All data produced in the present study are available upon reasonable request to the authors.

## Acknowledgement

This work was supported by Vanke School of Public Health, Tsinghua University.

## Author contributions

Q.D. contributed to the conceptualization and methodology of the study; H.L. and Q.D. performed the analyses and the original draft preparation; D.S., Z.Y., W.H., and Q.D. contributed to the review and editing of the manuscript.

## References

1. Tsubokura M, Gilmour S, Takahashi K, Oikawa T, Kanazawa Y. Internal Radiation Exposure After the Fukushima Nuclear Power Plant DisasterJAMA 2012; 308(7): 669.

2. Yamanishi T, Kakiuchi H, Tauchi H, Yamamoto T, Yamamoto I. Discussions on tritiated water treatment for Fukushima Daiichi nuclear power station. Fusion Science and Technology 2020; 76(4): 430–8.

3. Normile D. Fukushima wastewater release set to start soon. Science (New York, NY) 2023; 379(6630): 321–.

4. Lehto J, Koivula R, Leinonen H, Tusa E, Harjula R. Removal of Radionuclides from Fukushima Daiichi Waste Effluents. Separation &amp; Purification Reviews 2019; 48(2): 122–42.

5. TEPCO. Current ALPS Treated Water, etc. Conditions. 2023. https://www.tepco.co.jp/en/decommission/progress/watertreatment/alps01/index-e.html (accessed March 132023.

6. Zhao C, Wang G, Zhang M, et al. Transport and dispersion of tritium from the radioactive water of the Fukushima Daiichi nuclear plant. Marine pollution bulletin 2021; 169: 112515.

7. Liu Y, Guo X-Q, Li S-W, Zhang J-M, Hu Z-Z. Discharge of treated Fukushima nuclear accident contaminated water: macroscopic and microscopic simulations. National Science Review 2022; 9(1): nwab209.

8. Bezhenar R, Takata H, de With G, Maderich V. Planned release of contaminated water from the Fukushima storage tanks into the ocean: Simulation scenarios of radiological impact for aquatic biota and human from seafood consumption. Marine Pollution Bulletin 2021; 173: 112969.

9. Guo J, Liu Y, Wu X, Chen J. Assessment of the impact of fukushima nuclear wastewater discharge on the global economy based on GTAP. Ocean & Coastal Management 2022; 228: 106296.

10. Behrens E, Schwarzkopf FU, Lübbecke JF, Böning CW. Model simulations on the long-term dispersal of 137Cs released into the Pacific Ocean off Fukushima. Environmental Research Letters 2012; 7(3): 034004.

11. Lai Z, Chen C, Beardsley R, et al. Initial spread of 137Cs from the Fukushima Dai-ichi Nuclear Power Plant over the Japan continental shelf: a study using a highresolution, global-coastal nested ocean model. Biogeosciences 2013; 10(8): 5439–49.

12. Periáñez R, Qiao F, Zhao C, et al. Opening Fukushima floodgates: Modelling 137Cs impact in marine biota. Marine Pollution Bulletin 2021; 170: 112645.

13. Evangeliou N, Balkanski Y, Cozic A, Møller AP. Global and local cancer risks after the Fukushima Nuclear Power Plant accident as seen from Chernobyl: A modeling study for radiocaesium (134Cs & 137Cs). Environment international 2014; 64: 17–27.

14. Yamaguchi M. Fukushima plant failed to probe cause of faulty filters. PBS NEWS HOUR. 2021.

15. Jizhou Zhu ea. Nuclear reactor safety analysis: Xi’an Jiaotong University Press; 2000.

16. Japan Atomic Industrial Forum I. Information on Status of Nuclear Power Plants in Fukushima 2011.

17. NEMO TOP Working Group. Tracers in Ocean Paradigm (TOP) --The NEMO Tracers engine. Scientific Notes of Climate Modelling Center: Institut Pierre-Simon Laplace (IPSL); 2022.

18. Copplestone D, Bielby S, Jones S, Patton D, Daniel P, Gize I. Impact assessment of ionising radiation on wildlife. 2001.

19. Vives IBJ, Wilson RC, Watts SJ, Jones SR, McDonald P, Vives-Lynch S. Dynamic model for the assessment of radiological exposure to marine biota. J Environ Radioact 2008; 99(11): 1711–30.

20. IAEA. Sediment Distribution Coefficients and Concentration Factors for Biota in the Marine Environment.: IAEA 2004.

21. Williams J, Russ R, McCubbin D, Knowles J. An overview of tritium behaviour in the Severn Estuary (UK). Journal of radiological protection 2001; 21(4): 337.

22. Ministry of Health, Labour and Welfare, Japan. National Health and Nutrition Survey. 2019.

23. ICRP. Age-dependent Doses to the Members of the Public from Intake of Radionuclides - Part 5 Compilation of Ingestion and Inhalation Coefficients. ICRP Publication 72; 1995.

24. Moon E-K, Ha W-H, Seo S, et al. Estimates of Radiation Doses and Cancer Risk from Food Intake in Korea. Journal of Korean Medical Science 2016; 31(1): 9.

25. Valentin J. The 2007 recommendations of the international commission on radiological protection: Elsevier Oxford; 2007.

26. Independent NA. The Fukushima Nuclear Accident Independent Investigation Commission. The National Diet of Japan Tokyo, Japan; 2011.

27. Metivier H. Chernobyl, Assessment of Radiological and Health Impacts: Nuclear Energy Agency, Organisation for Economic Co-operation and Development; 2002.

28. Buesseler KO. Opening the floodgates at Fukushima. Science 2020; 369(6504): 621–2.

29. TEPCO. Draft Study Responding to the Subcommittee Report on Handling ALPS Treated Water 2020.

30. Cao Y, Zhao Z, Wang P, et al. Long-term variation of 90Sr and 137Cs in environmental and food samples around Qinshan nuclear power plant, China. Scientific Reports 2021; 11(1).

31. Kenyon JA, Buesseler KO, Casacuberta N, et al. Distribution and Evolution of Fukushima Dai-ichi derived 137Cs, 90Sr, and 129I in Surface Seawater off the Coast of Japan. Environmental science &technology 2020; 54(23): 15066–75.

32. Tazoe H, Yamagata T, Tsujita K, et al. Observation of Dispersion in the Japanese Coastal Area of Released 90Sr, 134Cs, and 137Cs from the Fukushima Daiichi Nuclear Power Plant to the Sea in 2013. International Journal of Environmental Research and Public Health 2019; 16(21): 4094.

33. Stohl A, Seibert P, Wotawa G, et al. Xenon-133 and caesium-137 releases into the atmosphere from the Fukushima Dai-ichi nuclear power plant: determination of the source term, atmospheric dispersion, and deposition. Atmospheric Chemistry and Physics 2012; 12(5): 2313–43.

34. Koo Y-H, Yang Y-S, Song K-W. Radioactivity release from the Fukushima accident and its consequences: A review. Progress in Nuclear Energy 2014; 74: 61–70.

35. Yasunari TJ, Stohl A, Hayano RS, Burkhart JF, Eckhardt S, Yasunari T. Cesium-137 deposition and contamination of Japanese soils due to the Fukushima nuclear accident. Proceedings of the National Academy of Sciences 2011; 108(49): 19530–4.

36. Casacuberta N, Masqué P, Garcia-Orellana J, Garcia-Tenorio R, Buesseler KO. 90Sr and 89Sr in seawater off Japan as a consequence of the Fukushima Dai-ichi nuclear accident. Biogeosciences 2013; 10(6): 3649–59.

37. Kuwata H, Akata N, Okada K, et al. Monthly Precipitation Collected at Hirosaki, Japan: Its Tritium Concentration and Chemical and Stable Isotope Compositions. Atmosphere 2022; 13(5): 848.

38. Koarashi J, Nishimura S, Atarashi-Andoh M, Muto K, Matsunaga T. A new perspective on the 137Cs retention mechanism in surface soils during the early stage after the Fukushima nuclear accident. Scientific Reports 2019; 9(1).

39. Harada KH, Niisoe T, Imanaka M, et al. Radiation dose rates now and in the future for residents neighboring restricted areas of the Fukushima Daiichi Nuclear Power Plant. Proceedings of the National Academy of Sciences 2014; 111(10): E914–E23.

40. Organization WH. Preliminary dose estimation from the nuclear accident after the 2011 Great East Japan Earthquake and Tsunami: World Health Organization; 2012.

41. Ten Hoeve JE, Jacobson MZ. Worldwide health effects of the Fukushima Daiichi nuclear accident. Energy &amp; Environmental Science 2012; 5(9): 8743.

42. Drozdovitch V, Bouville A, Chobanova N, et al. Radiation exposure to the population of Europe following the Chernobyl accident. Radiation Protection Dosimetry 2007; 123(4): 515–28.

43. Liu D, Hoskin M. Contemporary international Law: Regulating the upcoming fukushima radioactive wastewater discharge. Ocean &Coastal Management 2023; 234: 106452.

