## Supplementary Information for "Health impact of nuclear waste water discharge from the Fukushima Daiichi nuclear plant"

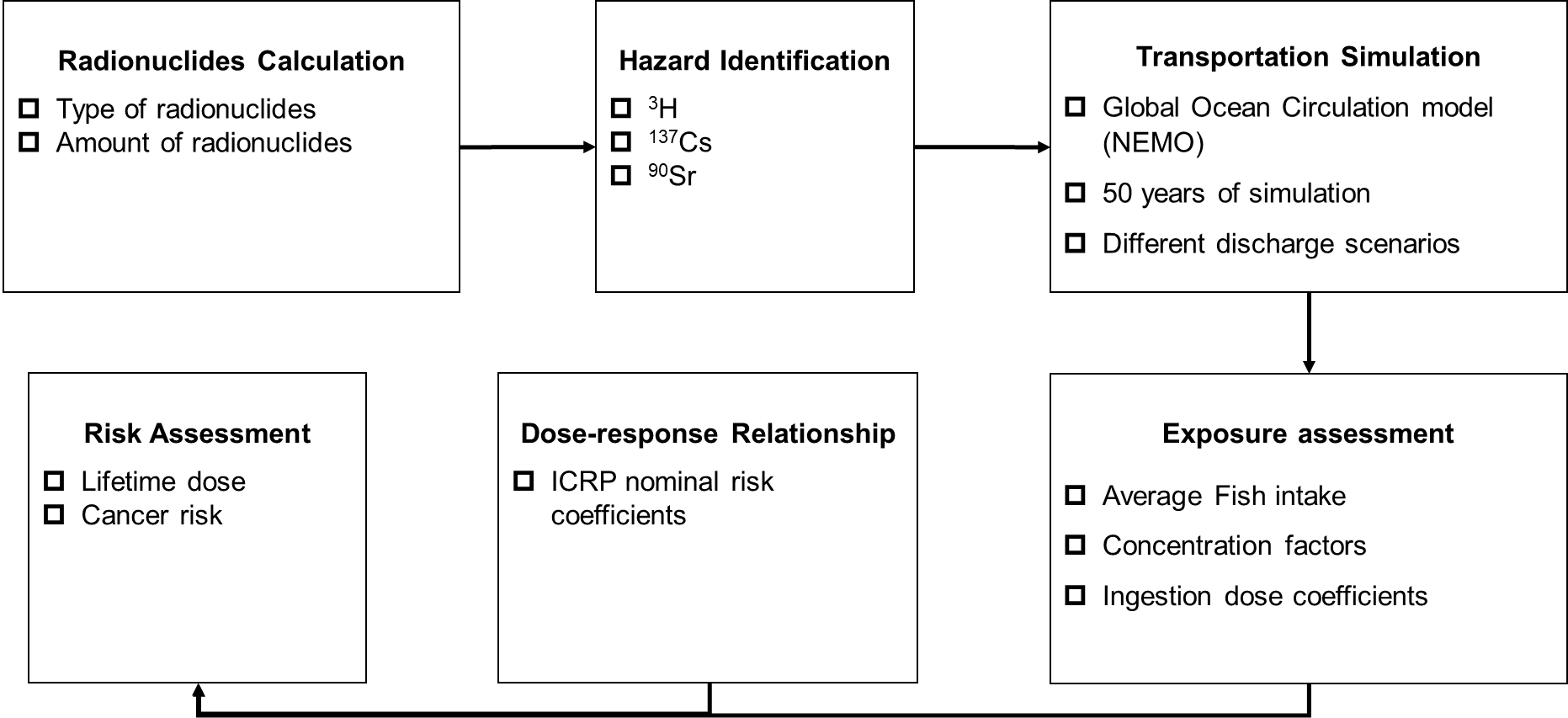

**Fig.S1 Study framework**

**Table S1 Important radioactive fission products**

| **Nuclide** | **Half-life** | **Activity** | **Radiation type** |
| --- | --- | --- | --- |
| ^131^I | 8.1d | 940 | β, γ |
| ^134^Cs | 2.1a | 140 | β, γ |
| ^137^Cs | 30.1a | 70 | β, γ |
| ^90^Sr | 30.2a | 52 | β |
| ^106^Ru | 1.0a | 310 | β |
| ^3^H | 12.3a | 1.42 | β |

I (iodine) isotopes emit high-energy beta and gamma rays, making them significant contributor to the external dose of radioactivity emitted from the dust. Additionally, iodine has a tendency to accumulate in the thyroid gland, leading to internal irradiation of the organ. The most critical iodine isotope, ^131^I, is often used a measure of the severity of the accident.

Cs (cesium) shares similar chemical properties with K (potassium). The chemical reaction between Cs and I will affect the release amount and chemical composition. Cesium is absorbed into the body through muscle tissue and then separated within a few months. This time is shorter than the half-life of ^137^Cs, so the amount of ^137^Cs in the body quickly balances out with the amount in food. Meat and milk are important ways for Cs to enter the body.

^90^Sr (strontium) and ^106^Ru (Ruthenium) emit only beta suspicion and are not easy to measure. Sr is volatile, but its oxides are not. The opposite is true for Ru. Therefore, the oxidation state in the reactor has a great influence on the release form of fission products. Sr enters the human body through milk, and the sensitive organ is bone, and elimination is slow.

**Table S2 Total amount of radionuclides after Fukushima Daiichi nuclear power plant accident in Condition 1**

| **Radionuclides** | **Half-life (a)** | **Activity concentration per unit (10^12^ Bq /MW)** | **Initial activity concentration (10^12^ Bq)** | **Activity concentration after 12 years (10^12^ Bq)** |
| --- | --- | --- | --- | --- |
| ^3^H | 12.3000 | 1.4200 | 4970.0000 | 2527.7315 |
| ^131^I | 0.0222 | 940.0000 | 3,290,000.0000 | - |
| ^134^Cs | 2.1000 | 140.0000 | 490,000.0000 | 9,340.9260 |
| ^137^Cs | 30.1000 | 70.0000 | 245,000.0000 | 185,857.3000 |
| ^90^Sr | 28.9000 | 52.0000 | 182,000.0000 | 138,191.8000 |
| ^106^Ru | 1.0000 | 310.0000 | 1,085,000.0000 | 265.3608 |
| ^238^Pu | 89.0000 | 1.3000 | 4,550.0000 | 4,144.1140 |
| ^239^Pu | 24,000.0000 | 0.2800 | 980.0000 | 979.6605 |
| ^240^Pu | 6,580.0000 | 0.3100 | 1,085.0000 | 1,083.6300 |
| ^241^Pu | 14.7000 | 5.6000 | 19,600.0000 | 11,131.9000 |
| ^242^Pu | 380,000.0000 | 0.0005 | 1.7500 | 1.74996 |
| ^242^Cm | 0.4500 | 15.0000 | 52,500.0000 | 0.0005 |
| ^244^Cm | 18.2000 | 0.9100 | 3,185.0000 | 2,016.8350 |

Note: For Condition 1, we calculated the upper limit amount of radioactive substances based on damage condition or cores and damaged power of reactors. More details were articulated in the Method Section. ^131^I would be rarely found after 12 years of decay.

**Table S3 Total amount of radionuclides after Fukushima Daiichi nuclear power plant accident in Condition 2**

| **Radionuclides** | **Half-life (a)** | **Activity concentration per unit (10^12^ Bq /MW)** | **Initial activity concentration (10^12^ Bq)** | **Activity concentration after 12 years (10^12^ Bq)** |
| --- | --- | --- | --- | --- |
| ^3^H | 12.3000 | 1.4200 | 11928.0000 | 6066.5557 |
| ^131^I | 0.0222 | 940.0000 | 7,896,000.0000 | - |
| ^134^Cs | 2.1000 | 140.0000 | 1,176,000.0000 | 22,418.2200 |
| ^137^Cs | 30.1000 | 70.0000 | 588,000.0000 | 446,057.5000 |
| ^90^Sr | 28.9000 | 52.0000 | 436,800.0000 | 331,660.3000 |
| ^106^Ru | 1.0000 | 310.0000 | 2,604,000.0000 | 636.8660 |
| ^238^Pu | 89.0000 | 1.3000 | 10,920.0000 | 9,945.8740 |
| ^239^Pu | 24,000.0000 | 0.2800 | 2,352.0000 | 2,351.1850 |
| ^240^Pu | 6,580.0000 | 0.3100 | 2,604.0000 | 2,600.7110 |
| ^241^Pu | 14.7000 | 5.6000 | 47,040.0000 | 26,716.5700 |
| ^242^Pu | 380,000.0000 | 0.0005 | 4.2000 | 4.19991 |
| ^242^Cm | 0.4500 | 15.0000 | 126,000.0000 | 0.0012 |
| ^244^Cm | 18.2000 | 0.9100 | 7,644.0000 | 4,840.4030 |

Note: The worst condition (referred as Condition 2 in the Method Section) assumed all 4 rectors and all cores were damaged; and the total damaged electric power was 2800 MWe. We calculated corresponding upper limit amounts of yielded radionuclides under this hypothetical scenario. ^131^I would be rarely found after 12 years due to short half-life.

**Video S1 Spatial dynamics of ^3^H under four discharge scenarios over the next 50 years**

**Video S2 Spatial dynamics of ^137^Cs under four discharge scenarios over the next 50 years**

**Video S3 Spatial dynamics of ^90^Sr under four discharge scenarios over the next 50 years**
